# Another step toward final call on Remdesivir efficacy as a treatment for hospitalized COVID-19 patients: a multicenter open-label trial

**DOI:** 10.1101/2021.08.13.21261992

**Authors:** Hamed Hosseini, Anahita Sadeghi, Payam Tabarsi, Azin Etemadimanesh, Ilad Alavi Darazam, Nasser Aghdami, Saeed Kalantari, Mehrdad Hasibi, Azar Hadadi, Farhang Babamahmoodi, Mansooreh Momen-Heravi, Ahmad Hormati, Yunes Panahi, Rozita Khodashahi, Mohammadreza Salehi

## Abstract

**Introduction:** After emerging the global pandemic of SARS-CoV2 some preliminary studies demonstrated the efficacy of antiviral treatments. But shortly thereafter, inconsistencies in the results of further clinical trials raised doubts on the efficacy of these agents. In this study, we aimed to evaluate the effect of Remdesivir on hospitalized COVID-19 patients’ outcomes.

**Material and methods:** This study was an open-label, single-armed, clinical trial on hospitalized patients diagnosed with COVID-19 who had progressive respiratory symptoms despite receiving standard care. All patients received Remdesivir and their characteristics, outcomes, time of treatment initiation, and respiratory support stages during hospitalization were registered and followed up for 14 days.

**Results:** 145 patients with the mean age of 52.89 ± 1.12 years enrolled in this study, 38 (26.2%) died at the end of 14 days period. The mean time interval from the onset of the symptoms to antiviral treatment was 10.63±0.56 days. Thirty deceased patients (78.9%) were men, showing 2.8 times higher mortality chance compared to women (OR_adj_=2.77; 95%CI=1.08-7.09). The type of respiratory support on the first day of treatment initiation showed a significantly lower mortality chance in patients receiving O_2_ only than those who needed non-invasive and/or mechanical ventilation (OR_adj_=3.91; 95%CI=1.64-9.32). The start time (early vs late administration) and duration (less or more than 7 days) of antiviral treatment had no statistically significant association with mortality or ventilation escalation among the patients (p-value > 0.05).

**Conclusion:** In this study, we showed that Remdesivir probably is not effective on the outcome of hospitalized COVID-19 patients.

## Introduction

The COVID-19 pandemic that began in late 2019, has impacted most parts of the world and posed major challenges to global health.[1] Approximately 105 million confirmed cases and 2.3 million COVID-19-related deaths were reported globally as of February 2021. In past year Iran reported more than 1.4 million confirmed COVID-19 cases.[2] In a published report of the Iranian COVID-19 national registry, the mortality rate of COVID-19 hospitalized patients was estimated to be 24.4% (23.8-25.0, 95% CI).[3]

Since the start of the pandemic, numerous treatment protocols have been formulated and tested around the world and, as of now, none of them has been shown to have significant effects on the mortality rate of COVID-19 [4].

Remdesivir, GS-5734, was one of the proposed antiviral drugs in COVID-19 treatment which was shown to be effective for the first time in a preliminary clinical trial and soon after World Health Organization (WHO) and Food and Drug Administration (FDA) approved and suggested it for emergent use in the treatment of COVID-19 [5-7].

While much more evidence was required to ensure the efficacy of Remdesivir, it was widely integrated with the COVID-19 treatment protocols worldwide. The dosage, indication of beginning and length of treatment and varies throughout the world[8].

In the meantime, the first multicenter randomized clinical trial of Remdesivir on 237 patients in China showed that it has no significant effect on the clinical recovery duration and the mortality rate of severe cases [9]. Controversies in the outcome of further clinical trials raised concerns about its effectiveness[8].

The most recent report of Solidarity megatrial on over 11 million hospitalized COVID-19 patients, measured the impact of suggested drugs on three main outcomes for COVID-19 patients; death, need for assisted ventilation, and length of hospitalization. The last published report stated that these main outcomes were not significantly reduced by none of the study drugs, including Remdesivir [10].

Expenditures and potential side effects caused by the widespread use of Remdesivir suggested the need for further clinical trials to settle the debate over its cost-effectiveness. For this purpose, in this trial we evaluated the efficacy of Remdesivir in Iranian hospitalized COVID-19 patients.

## Material and methods

This study was an open-label, single-armed, multicenter clinical trial performed on moderate to severe COVID-19 adult and non-pregnant hospitalized patients from 2020 March 20^th^ to 2020 July 14^th^ at 11 centers in 7 provinces of Iran. SARS-CoV2 infection was previously confirmed in all patients by nasopharyngeal sample SARS-CoV2 RT-PCR. Inclusion criteria included the need for hospitalization based on national COVID-19 protocol of Iran Health Ministry, progressive respiratory symptoms (Oxygen saturation (SpO2) dropped lower than 85% and/or tachypnea (RR>30 bpm) in room air) despite receiving standard care including hydration, Atazanavir/ritonavir or Lopinavir/ritonavir and O2 supplementation for at least 72 hours. None of the individuals had a serious life-threatening comorbidity include kidney, heart and liver failure and malignancies. Informed consent was obtained from each patient or his/her guardian beforehand, and the clinical trial protocol was approved by local ethics committee (code: IR.AJUMS.REC.1399.407) and registered in the Iranian Registry of Clinical Trials (code: IRCT20200404046937N5). In addition to standard treatment, all of the participants received 200mg intravenous Remdesivir on the first day and 100 mg daily for at least 4 days (with a minimum and maximum length of 5 and 10 days, respectively), and subsequently, they were monitored closely for 14 days from Remdesivir treatment initiation.

Collected data included the demographics, symptoms at the baseline, type and escalating of needed respiratory supports, and their outcome (discharged or expired, with date and cause of death).

Patients respiratory status was evaluated and registered every day based on a scored scale from 1 to 5 (1= discharged from the hospital, 2= hospitalized but no need for excessive O2 support, 3= O2 support with face mask/nasal cannula (O_2_ only), 4= receiving non-invasive ventilation/high-flow nasal cannula, 5= invasive mechanical ventilation). The first (on the first day of Remdesivir administration) and the last (within the first 14 days) respiratory status scores were compared between the groups, and possible improvements within and between the groups were investigated. The last respiratory status score was either recorded on the 14th day or before the patient’s discharge or death.

Patients outcomes were analyzed based on the duration from first symptoms to Remdesivir administration (less than 7 days or more than 7 days), type of needed respiratory support at the baseline of antiviral treatment initiation, and its changes and during follow up days.

No sample-size calculation was performed. All of the eligible patients who was independently volunteered to participate in this trial were included in the study based on drug inventory at the time. Trained research staff in each center recorded patients’ data on printed forms. Ultimately, data were analyzed using IBM SPSS© statistics 20.0. Mean (standard deviation) was used to describe quantitative data and frequency (percentage) was used for discrete data. Comparison of patients’ subgroups was performed with Chi-square and t-test. The univariate analysis was performed and variables with significant relevance (P-value>0.2) were selected as potential predictors of death. The logistic regression model was used to estimate the potential predictors of death in hospitalized patients. All statistical tests were reported at a 95% confidence level.

## Results

During this period, 263 cases were evaluated and 190 patients became candidates for inclusion in the study. Forty-five patients refused to fill out the consent form. By the end of the enrollment, a total number of 145 patients had been included in this clinical trial. (Figure 1) The mean age of participants was 52.89 ± 1.12 years (17-89 years). Fifty-one patients (35.2%) were female and the most frequent underlying diseases were diabetes (26.8%) and hypertension (23.4%). The mean and median time from the onset of symptoms to hospitalization were 7.17±0.47 and 6 days, respectively (min-max: 1-37 days).

**Figure 1.**
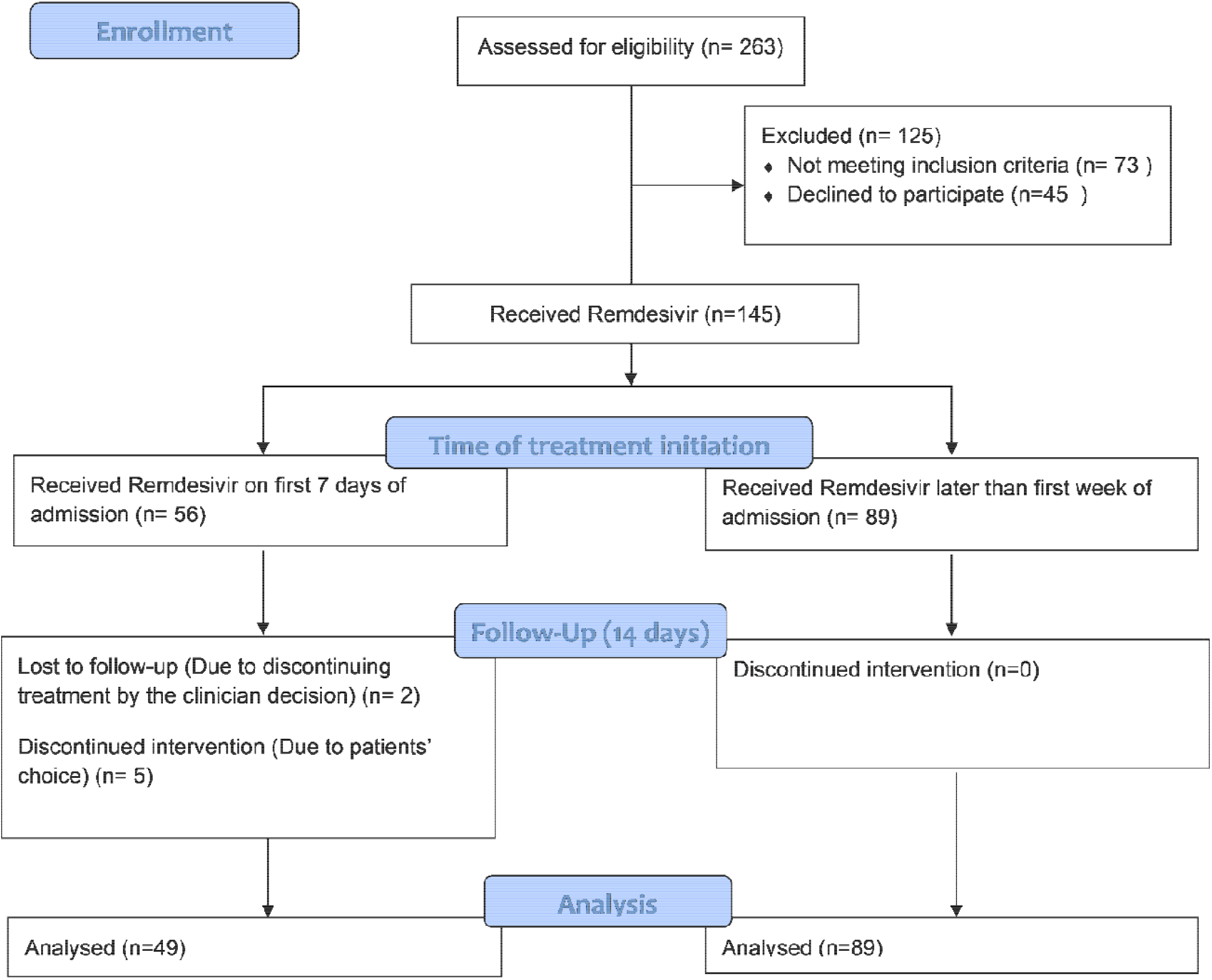
CONSORT flow chart of clinical trial

Furthermore, the mean time interval from the onset of the symptoms to receiving the first dose of Remdesivir among the deceased and discharged patients was comparable (10.63±5.16 and 10.70±7.0 days, respectively).

Out of 145 patients treated with Remdesivir, 38 (26.2%) died and 100 (69%) were discharged from the hospital without the need for further ventilatory support, among them 20 patients needed hospitalization more than 2 weeks. 7 (4.8%) patients abandoned the study during hospitalization. The median of length of hospitalization among survived patients was 7 days (IQR=8). Among the patients who died, the mean duration from the onset of the symptoms to death was 11.28 days (95%CI: 9.39-13.20). The mean age of the survivors was 53±14.38, and the mean age of deceased patients was 52.64±12.77 (p-value = 0.64). Of deceased patients, 30 (78.9%) were male and 8 (21.1%) were female (p-value = 0.047). (Table.1)

**Table 1.** Comparison of patients’ characteristics based on outcome at the end of study period (14 days)

| Variable | Value(s) for |  |  | P.value |
| --- | --- | --- | --- | --- |
|  | Total<br>(n=138) | survived<br>N=100 | mortality<br>group<br>N=38 | Live<br>vs.<br>Dead |
| Age, mean ±SD | 52.89 ± 1.12 | 58.84±14.38 | 52.64±12.77 | 0.64 |
| Male, n (%) | 91 | 61(67.0%) | 30(33.0%) | 0.05 |
| receiving IMV/NIV/HFNC at the<br>baseline | 74 | 45(60.8%) | 29(39.2%) | 0.001 |
| Duration, in days, of symptoms<br>before admission (mean SD) | 7.17±0.46 | 6.17±4.14 | 7.53±5.97 | 0.21 |
| Time from starting symptoms to start<br>Remdesivir (means SD) | 10.63±0.56 | 10.64±5.17 | 10.70±7.01 | 0.96 |
| >7 days from onset of symptoms to<br>receiving Remdesivir, n (%) | 89(64.5%) | 64(64%) | 25(65.8%) | 0.76 |

Multivariate analysis of the ventilatory support at the baseline showed that the patients on IMV / NIV / HFNC at the base time of receiving Remdesivir had about four times higher mortality chance compared with the patients receiving O_2_ only (OR_adj_=3.91; 95%CI=1.64-9.32). (Figure.2) Analysis of patients’ respiratory support status scores at the baseline demonstrated that increasing one score raises the mortality chance by 3.74 times (p-value= 0.008). Besides, mortality among men was 2.8 times more than women (OR_adj_=2.77; 95%CI=1.08-7.09). While the type of baseline respiratory support and gender were the only predictive variables for the outcome of hospitalized COVID-19 patients, other variables including length of receiving Remdesivir, time from starting symptoms to treatment initiation, age, and underlying diseases had no significant difference between the survived and the deceased group.

**Figure 2.**
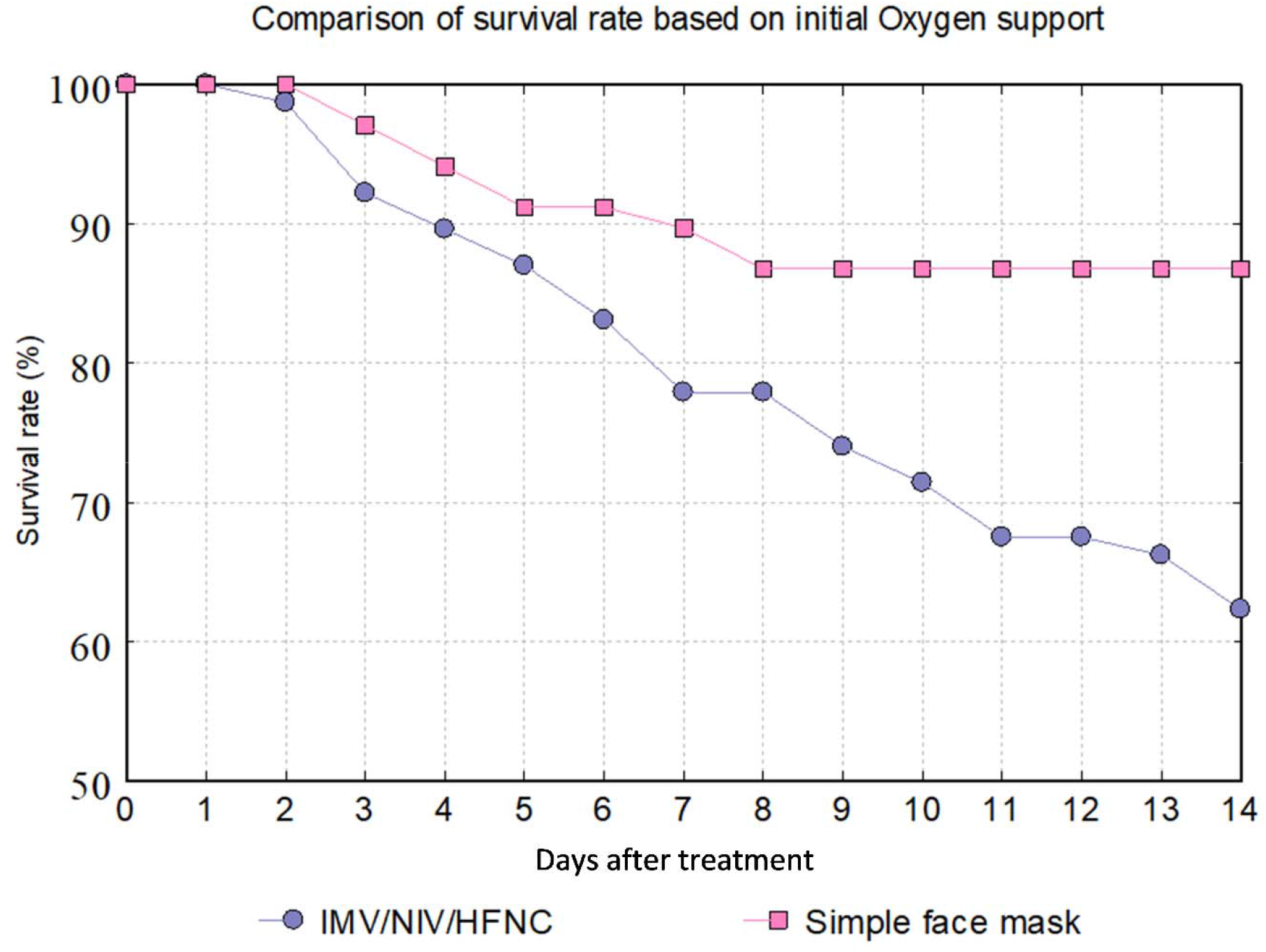
Day by day cumulative survival rate (%) based on initial Oxygen support compared between patients receiving Oxygen support with simple face mask and group that needed advanced Oxygen support with IMV, NIV or HFNC, IMV: invasive mechanical ventilation, NIV: non-invasive ventilation, HFNC: high-flow nasal cannula, IMV: Intermittent mandatory ventilation

The patients’ respiratory status score at the baseline (treatment initiation) and the end of the study (before death or on the 14th day) was evaluated and compared (Figure.3). The patients receiving mechanical ventilation and the ones who received only O2 support at the baseline had the lowest (20%) and the highest (61.6%) improvement in their respiratory status, respectively.

**Figure 3.**
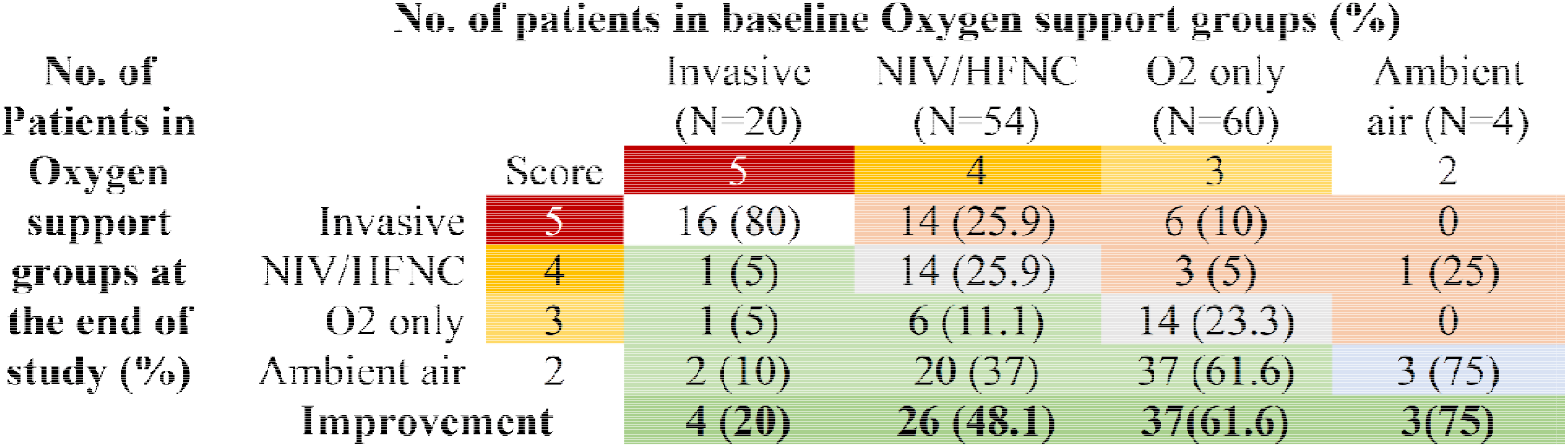
Oxygen support changes from the baseline to death or last day of study after treatment. Improvement (green), no change (gray), and worsening (pink) in oxygen status score are shown.

The demographics and characteristics of the participants were also compared between the patients receiving Oxygen only and the patients receiving IMV/NIV/HFNC at the initiation of the treatment (Table.2). The mean age of the patients receiving Oxygen via O2 only and those who received IMV/NIV/HFNC were 50.40±11.91 and 55.14±14.25, respectively (p-value = 0.04).

**Table 2.** Comparison of characteristics based on Oxygen support type at the baseline of antiviral treatment initiation

| Variable | Value(s) for |  | P.value |
| --- | --- | --- | --- |
|  | O<sub>2</sub> only | IMV/NIV/HFNC |  |
|  | n=67 | n=77 |  |
| Male, n (%) | 43(35.8%) | 51(66.2%) | 0.8 |
| Duration, in days, of symptoms before admission (mean ±SD) | 7.58±6.03 | 6.81±5.04 | 0.42 |
| Time from starting symptoms to start Remdesivir (means ±SD) | 9.89±7.27 | 11.28±5.80 | 0.22 |
| > 7 days from onset of symptoms to receiving Remdesivir, n (%) | 35(55.6%) | 55(76.4%) | 0.01 |
| Mortality, n (%) | 9(23.7%) | 29(76.3%) | 0.001 |

No serious adverse events related to antiviral therapy were observed in patients during the study. Remdesivir administration was discontinued for one patient by clinicians’ decision due to the raise in the liver enzymes, However, no association with drug administration was confirmed. Another patient refused to continue the treatment on second day of administration because of ‘not feeling good’, despite all good clinical of laboratory findings.

## Discussion

Months after the onset of the novel coronavirus pandemic, casualties of SARS-CoV2 infection and COVID-19-related pneumonia are still on the rise, however, there is as yet no established and efficient treatment protocol for these patients. [11]. As a nucleoside analog antiviral, Remdesivir was suggested to be a probable effective agent for treating COVID-19 based on its in-vitro and in-vivo effects on RNA-virus infections like the Ebola virus and SARS-CoV [12]. Some experimental and preliminary studies have shown that Remdesivir is an effective antiviral against SARS-CoV2, and since then, it has been commonly used as a choice for COVID-19. However, shortly afterward, Subsequent studies have reinforced this notion and have shown that Remdesivir has no significant impact on the outcome of the patients in need of mechanical ventilation, and only short-term effects in early administration have been observed [13]. On the other hand, some other recent trials have shown evidence of drug efficacy in the patients’ outcomes [14].

Results of this study showed 27.5% mortality rate among hospitalized COVID-19 patients receiving Remdesivir. Outcome of treatment was not related to the time and length of antiviral administration. Although lack of control group and randomization that makes the results of this study non-generalizable, adding up these findings to results of bigger studies may give us a clearer view on low efficacy of Remdesivir administration on outcome of hospitalized COVID-19 patients.

Among evaluated variables in this study biological gender can be a determining factor in predicting the outcome of the patients with a 2.8 times higher mortality chance in men. This difference between the two genders has already been observed in other studies, and it is unclear if the difference in the prevalence of underlying diseases between the two genders has a role in this finding or not. Further investigations will hopefully shed more light on this issue [15].

Another predictive variable in this study was the type of needed ventilatory support at the beginning of the antiviral treatment. This factor somehow represents the patients’ respiratory condition at the time, and since the acute respiratory distress syndrome (ARDS) is the main lethal complication of COVID-19, it can be considered as a sign of disease severity in patients [16]. Our results demonstrated that the need for advanced ventilatory support like NIV, IMV, and HFNC on the Remdesivir administration day raises the mortality chance four times compared with patients who received O2 only. This finding is also mentioned in some other clinical trials on severe COVID-19 patients that showed receiving Remdesivir shortened the time to recovery interval in early administration but it had no significant benefit on the clinical outcome of the patients [9]. Also, there was a correlation between age and the need for advanced O2 support which is assumed to be a predictive value for COVID-19 outcome.

The mortality rate of COVID-19 among about 29000 Iranian patients was reported on April 2020 by the Iranian registry of COVID-19. This report consisted of divided mortality rates in different subgroups with various comorbidities, like diabetes and cardiovascular diseases. A visual comparison of mortality rates in the current study, patients receiving Remdesivir, and general population treated with routine treatment protocol of Iran health ministry before integrating Remdesivir in the protocols, is presented in Figure 4.

**Figure 4.**
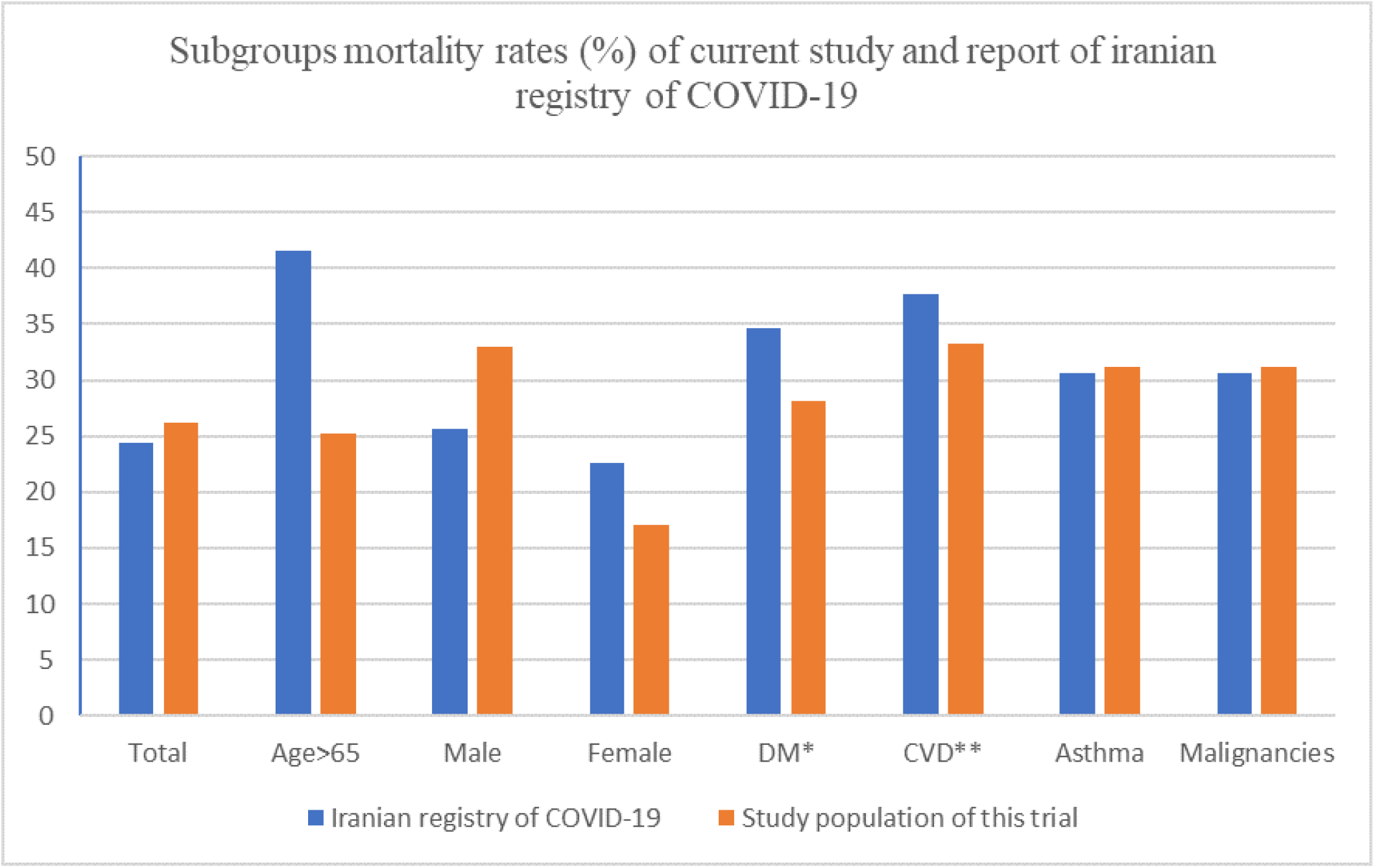
Comparison of mortality rate among Iranian COVID-19 registry (3) and Patients receiving Remdesivir in current trial, *Diabetes mellitus, **Cardiovascular diseases

We also found that between the deceased and living patients, Remdesivir administration within the first seven days after the onset of the symptoms or after that is not significantly different. Lack of significant difference in drug initiation time between the survived and deceased groups seemingly disproves the speculations about greater efficacy of Remdesivir in case of earlier treatment initiation and could highlight the negligible impact of this drug throughout the entire disease course. Based on the results of this study and similar trials, we can conclude that the timely initiation of Remdesivir is merely effective in the early stages of respiratory complications, and does not necessarily impact the survival rate in progression to critical complications.

In conclusion, the results of this study showed that Remdesivir probably is not effective in the outcome of hospitalized COVID-19 patients, especially in patients with more severe respiratory complications and we should keep trying to find more promising treatments for these patients.

## Data Availability

All data are available by request contacting the corresponding author

## Acknowledgments

Our sincere thanks to Professor Reza Malekzadeh who coordinated and supervised the project. We express our gratitude to the Digestive Diseases Research Institute (DDRI), Tehran University of Medical Sciences. Conducting the study would not have been possible without the efforts of the DDRI team. The authors also would like to express their gratitude for all care workers and patients in study hospitals. This study has been funded and supported by Tehran University of Medical Sciences (TUMS); Grant no. 99-1-97-47143.

